# A systematic review and meta-analysis on the rate of human schistosomiasis reinfection

**DOI:** 10.1101/2020.07.18.20156703

**Authors:** Abdallah Zacharia, Vivian Mushi, Twilumba Makene

**Affiliations:** Department of Parasitology and Medical Entomology, Muhimbili University of Health and Allied Sciences, Dar es Salaam, Tanzania

**Author notes:** **Corresponding author** (AZ).

**Keywords:** Human schistosomiasis, reinfection, Schistosoma mansoni, Schistosoma haematobium, Schistosoma japonicum

## Abstract

**Background:** While praziquantel mass drug administration is currently the most widely used method in the control of human schistosomiasis, it does not prevent subsequent reinfection hence persistent transmission. Towards schistosomiasis elimination, understanding the reinfection rate is crucial in planning for the future interventions. However, there is scarcity information on the global schistosomiasis reinfection rate.

**Objective:** This systematic review and meta-analysis aimed at summarizing studies that estimated the human schistosomiasis reinfection rate.

**Materials and methods:** The protocol for this review was prepared to guide authors throughout the process. However, registration was not sought. Three data bases (PubMed, Hinari and Google Scholar) were thoroughly searched to retrieve original research articles presenting data on human schistosomiasis reinfection rate. Study quality and risk of bias was assessed based on Joanna Briggs Institute critical appraisal checklist. Meta-analysis was conducted using statistical R version 3.6.2 and R Studio using “meta” and “metafor” packages. Heterogeneity and publication bias of the studies were determined using Q – test and Egger’s regression test respectively. Random effect model was employed to estimate pooled reinfection rates.

**Results:** A total of 29 studies met inclusion criteria to be included in this review. All studies had at least satisfactory (5 – 9 scores) quality. The overal calculated and pooled schistosomiasis reinfection rates were 36.1% (±23.3%) and 33.2% (95% CI, 26.5 – 40.5%) respectively. For intestinal schistosomiasis, the calculated and pooled reinfection rates were 43.9% (±20.6%) and 43.4% (95% CI, 35.8 – 51.4%), and that for urogenital schistosomiasis were 17.6% (±10.8%) and 19.4% (95% CI, 12.3% – 29.2%) respectively. Results of subgroup analysis showed that, the type of *Schistosoma* species, participants age group, sample size and geographical area had influence on disparity variation in Schistosomiasis reinfection rate (p < 0.05).

**Conclusion:** Despite the control measures in place, the re-infection rate was still high, specifically on intestinal schistosomiasis as compared to urogenital schistosomiasis. Achieving 2030 sustainable development goal 3 on good health and wellbeing through schistosomiasis elimination and intensive programmatic strategies should be implemented. Among such strategies to be used at national level are repeated mass drug administration at least every six month, intensive snails control and health education.

## 1 Introduction

Human schistosomiasis is one of the neglected tropical diseases caused by trematodes of the genus *Schistosoma*. Human schistosomiasis occur in two forms known as intestinal schistosomiasis and urogenital schistosomiasis (1). The main species causing intestinal schistosomiasis are; *Schistosoma mansoni, Schistosoma japonicum, Schistosoma mekongi and Schistosoma intercalatum* while urogenital schistosomiasis is caused by *Schistosoma haematobium* (2). These species require fresh water snail for the development of an infective stage of the parasite which afterward infect people as they come into contact with water. Human acquire the infection during routine domestic, occupational, agricultural and recreational activities which expose them to infested water (3). The groups at higher risk of acquiring schistosomiasis are pre-school aged children, school-aged children and people with occupations that involve contact with infested water such as irrigation workers, fishermen, farmers and women doing domestic chores in infested water (4).

The disease is more prevalent in tropical and subtropical countries particularly in poor communities where sanitation and water resources are of poor quality and inadequate (4,5). Poor water, sanitation and hygiene conditions such as the inadequate supply of clean and safe water, inadequate sanitation and unhygienic practices such as open defecation and urination play an important role in contamination of fresh water sources and therefore facilitate transmission of human schistosomiasis (6,7)

Human schistosomiasis lead to hepatomegaly, splenomegaly, anaemia, kidney malfunction, stunting growth in children and reduce ability to learn in school/cognitive dysfunction in children (8,9) The magnitude of human schistosomiasis varies across different areas with respect to time. In Sub-Saharan Africa alone, it was estimated that 436 million people in 78 countries live in endemic areas that put them at risk of urogenital schistosomiasis and over 112 million people were infected. For the case of intestinal schistosomiasis 393 million people were at the risk of infection and 54 million people were infected with more than 200,000 deaths (10).

The interventions such as preventive chemotherapy (praziquantel), snail control, health education and improvement of water supply and sanitation facilities have been going on in order to prevent and control schistosomiasis (11,12). Large scale use of preventive chemotherapy has been very useful in the reduction of morbidity and mortality due to schistosomiasis. However, schistosomiasis transmission is not yet interrupted by the current control measures (7,12). It has also been observed that treatment does not avert subsequent infection; thus, if contact with infested water is continued, re-infection can take place relatively quickly (13). Several factors have been associated with the rapid schistosomiasis reinfection rate such as age, gender, sex, pre-treatment intensity, level of immunity and water contact behaviour attributed to economic, domestic and recreational activities (14–16). However the effects of these factors were shown to vary depending on the type of *Schistosoma* species and its geographical distribution. Studies on the schistosomiasis reinfection rate have been documented in several parts of the world. Despite the available information on the factors contributing to the rapid schistosomiasis reinfection rate, to our knowledge, there is no review which described the schistosomiasis reinfection rate based on the evidence gathered from different parts of the world. The available article reviewed data on *Schistosoma japonicum* reinfection rate only in China (16). Hence, this study was designed to conduct a systematic review, with meta-analysis, of studies that estimated human schistosomiasis reinfection rate globally. A clear understanding of the schistosomiasis reinfection rate is crucial for planning effective and sustainable strategies to control schistosomiasis transmission.

## 2 Materials and methods

### 2.1 Search strategy

The protocol for this systematic and meta-analysis was prepared to guide authors throughout the process. However, registration was not sought. We conducted a thoroughly literature search of the following electronic databases; PUBMED, HINARI and Google Scholar. To obtain study articles containing information of our interest, the words “Schistosomiasis”, “*Schistosoma mansoni*”, “*Schistosoma haematobium*”, “*Schistosoma japonicum*” and “reinfection” joined using “OR” and “AND” booleans were used to create search query. We limited our search to journal articles only. There was no limitation of language of publication, year of publication and area where study was conducted. Search results from the three databases were combined together for selection process. The bibliographies of the selected articles were manually searched for the presence of other review related articles.

### 2.2 Study eligibility criteria

We included all studies presenting original research work with the following characteristics. Population: articles must have presented original research work conducted on human participants of any age, sex, race and from any geographical area. Intervention: the articles must have indicated that before follow-up, the study participants were diagnosed with any of human *Schistosoma* species and treated till cured. However there was no limitation on the type of drug, number of doses used and time interval from treatment to cure. Outcome: our outcome of interest was human schistosomiasis reinfection rate. The included articles must have presented data on the reinfection rate with any *Schistosoma* species. According to our definition, reinfection rate means the proportion of egg-positive participants who get cured (egg-negative by microscopic techniques) and then become egg-positive after a certain specified period of time. We excluded articles on experimental and non human studies, review articles, letter to editors and articles which did not report on reinfection rate as per our definition.

### 2.3 Study selection and data extraction

After combining search results from the three databases, we removed all duplicates. Reviewer AZ screened all searched articles based on the titles and abstracts for their eligibility to be included in full text review. Articles eligible for full text review were retrieved. All accessed full text articles were reviewed by two independent reviewers (AZ and VM) for the eligibility to be included in data extraction. Matched articles selected by the two reviewers were subjected to data extraction; whenever there was mismatch the third reviewer (TM) was involved. Data were independently extracted by the same two reviewers (AZ and VM) using a designed data extraction form. Prepared data extraction form was pre-tested by three reviewers before it had been used. When there was difference in data extracted by the two reviewers from similar article, the third reviewer (TM) was invited to independently extract the data. Similar data extracted by either one of the first two reviewers and the third reviewer was taken. When the third reviewer came with different data, consensus was reached through discussion. Extracted data included publication details (first author’s name, journal name and year of publication), methodology (study design, country of study, year of data collection, description of study setting, characteristics of study population, diagnostic test, drug(s) and dose(s) used, and follow-up time) and results (identified *Schistosoma* species and reinfection rate)

### 2.4 Assessment of study quality and risk of bias

The study quality and risk of bias for selected studies were assessed based on 9 criteria as described in the Joanna Briggs Institute critical appraisal checklist for use in reviews of prevalence studies (17). Each criterion was given either of the two options of YES if a criterion was met or NO if a criterion was not met. A YES option was graded as 1 and NO option as 0. The minimum score of 0 was given if all criteria were not met and maximum score of 9 was given if all criteria were met. Studies with overall grades ranging from 0 - 4 were considered of low quality, 5 - 7 moderate quality and 8 – 9 high quality. Studies with moderate to high quality were included in the review. Two reviewers (AZ and VM) independently assessed the quality of selected articles.

### 2.5 Data analysis

We used Statistical Package for Social Sciences version 24 (IBM Corp., Armonk, NY, USA) and statistical R version 3.6.2 (R Studio using “meta” and “metafor” packages) to perform descriptive statistical tests and meta-analysis respectively. The random effect model was used to calculate both the overall and subgroup pooled schistosomiasis reinfection rates.

#### 2.5.1 Heterogeneity and publication bias

The degree of heterogeneity was calculated using the restricted maximum likelihood method. Visual inspection of forest plots was used to assess any indicator of the presences of heterogeneity. We employed Q-test to identify the presence of any significant heterogeneity (significant at p □ 0.1); *r*^*2*^ *-* test was used for an estimation of between-studies variability; *I*^*2*^ – test was used to estimate the proportion of the observed between-studies variability that is due to real differences and not due to chances. *I*^*2*^ value above 50% was considered to imply significant heterogeneity. Publication bias was evaluated using funnel plot and confirmed using Egger’s regression analysis.

#### 2.5.2 Subgroup analysis

Subgroup analysis was performed to assess the effect of different factors on the pooled schistosomiasis reinfection rate. Factors assessed were; type of *Schistosoma species*, time to follow-up, study setting, sample size, geographical area and age groups. We could not include type of drug used as very few studies have not used praziquantel during treatment. These factors were categorized as follows; Schistosoma species into three categories (*Schistosoma haematobium, Schistosoma mansoni* and *Schistosoma japonicum*), follow-up time into two categories (less than 12 months and 12 months and above), study setting into two categories (school based studies and community based studies), geographical area into two categories (Africa and Non Africa) and age groups into two categories (less than 16 years and all ages), sample size (small; n < 100, moderate; n = 100 to 499 and large; n ≥ 500), age groups into two categories (less than 15 years and all ages).

## 3 Results

### 3.1 Search results and study selection

A total of 1045 search results were obtained from the three electronic databases. From these search results 145 duplicates were removed. Based on the primary screening of titles and abstracts, a total of 66 studies were eligible for full text review. Based on inclusion and exclusion criteria, 29 studies were found eligible for data synthesis process. Figure 1 is a flow diagram showing the stages followed during studies selection process.

**Fig 1.**
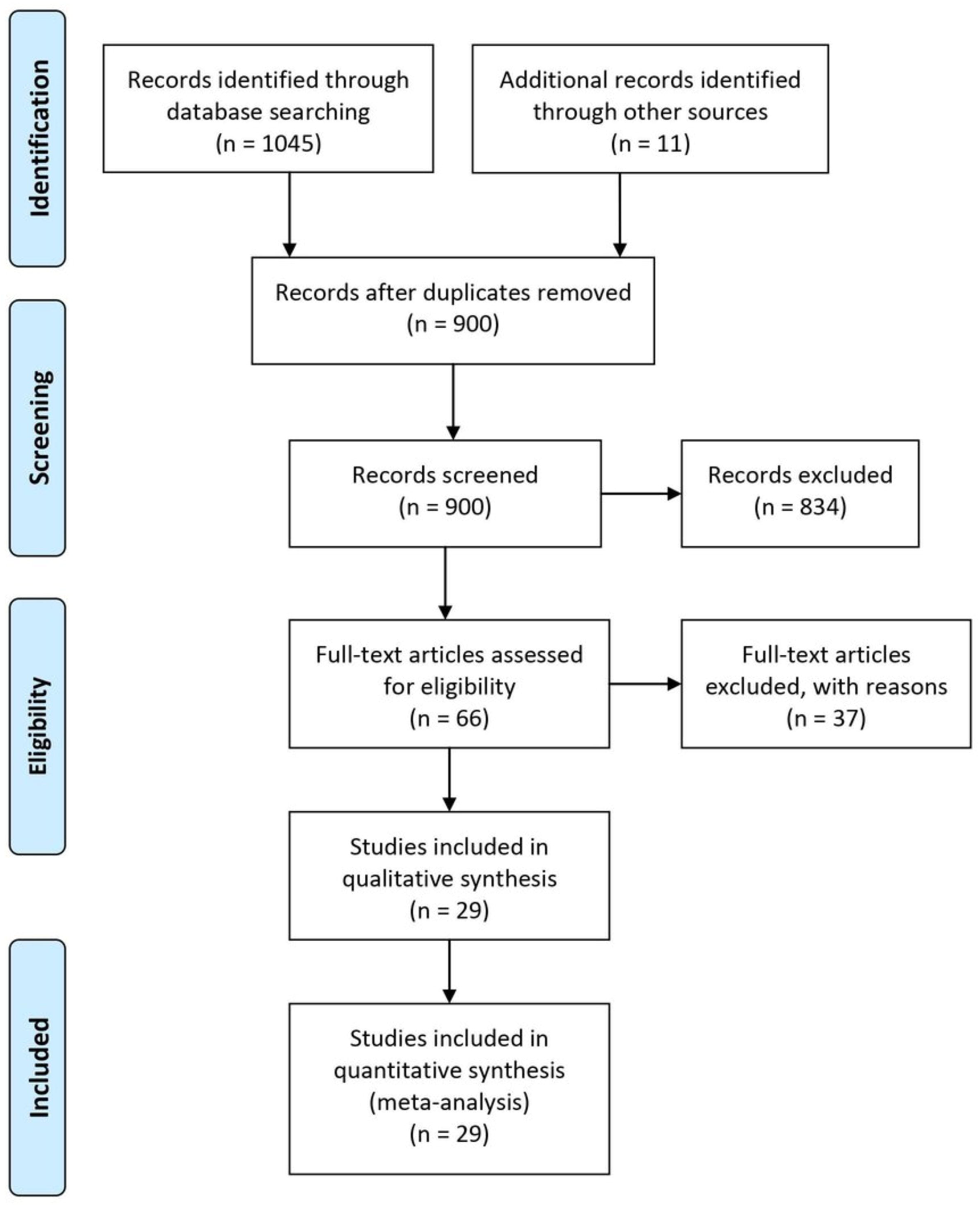
Flow diagram showing studies selection process

### 3.2 Study quality and risk of bias assessment results

Study quality and risk of bias assessment results showed no low quality study, 9 studies scored 5 – 7 (moderate quality) and 20 studies scored 8 - 9 (high quality). The average score was 7.9 which indicate the overall moderate quality of included studies. According to the criteria used 21 (72.4%) studies had adequate sample sizes. All studies used valid laboratory diagnostic tests (Kato-katz and Urine filtration) for the diagnosis of schistosomiasis.

### 3.3 Schistosomiasis reinfection

#### 3.3.1 Studies characteristics

Twenty nine studies published between the year 1991 and 2019 were included for data synthesis. Out of 29 included studies 54 datasets on Schistosomiasis reinfection rate caused by three *Schistosoma* species were extracted. Of 29 studies, 10 studies reported on *Schistosoma mansoni*, 12 studies reported on *Schistosoma haematobium*, 4 studies reported on *Schistosoma japonicum*, 2 studies reported on *Schistosoma mansoni* and *Schistosoma haematobium* and 1 study reported on *Schistosoma mansoni* and *Schistosoma japonicum*. These studies were conducted in 14 countries. Eleven out of 14 countries are in Africa, 2/14 countries are in Asia and 1/14 countries is in South America. The follow-up time ranged from 3 to 156 months. The age of participants ranged from 1 to 67 years with all of studies involved both male and female populations. Praziquantel and Oxamniquine antischistosoma drugs were used to cure study participants prior to follow-up in 27 and 2 studies respectively (Table 1).

**Table 1.** Characteristics of presented studies

| Study | Country | Study design | Study Setting | Age (years) | Diagnostic Test | Treatment Drug | Follow up (months) |
| --- | --- | --- | --- | --- | --- | --- | --- |
| ***Schistosoma mansoni*** |  |  |  |  |  |  |  |
| de Lima e Costa *et al.*, (18) | Brazil | PC | Community | ≥5 | Kato-katz | Oxamniquine | 156 |
| de Moira *et al.*, (19) | Uganda | Longitudinal | Community | 5 to 66 | Kato-katz | Praziquantel | 12 |
| Egesa *et al.*, (20) | Uganda | BF (no control) | Community | 6 to 40 | Kato katz | Praziquantel | 12 |
| Favre *et al.*, (21) | Brazil | RCT | SC | 6 to 15 | Kato katz | Praziquantel | 12 |
| Garba *et al.*, (22) | Niger | LC | Children | 6 to15 | Kato katz | Praziquantel | 6 and 12 |
| Gazzinelli *et al.*, (23) | Brazil | Longitudinal | Children | 6 to 15 | Kato-katz | Praziquantel | 12 |
| Munishi *et al.*, (24) | Tanzania | Longitudinal | SC | 6 to 16 | Kato katz | Praziquantel | 5 and 8 |
| Nalungwa *et al.*, (25) | Uganda | RCT | PSAC | 1 to 5 | Kato katz | Praziquantel | 8 |
| Olliaro *et al.*, (26) | Tanzania & Brazil | RCT | Children | 10 to 19 | Kato katz | Praziquantel | 6 and 12 |
| Reis *et al.*, (27) | Brazil | RCT | Children | 7 to 18 | Kato-katz | Oxamniquine | 6 |
| Satti *et al.*, (28) | Sudan | Longitudinal | Farmers | 25 to 55 | Kato-katz | Praziquantel | 12 |
| Woldegerima *et al.*, (29) | Ethiopia | Follow-up | SC | 9 to 14 | Kato katz | Praziquantel | 6 |
| ***Schistosoma haematobium*** |  |  |  |  |  |  |  |
| Chandiwana *et al.*, (30) | Zimbabwe | Cohort | Community | ≥2 | Filtration | Praziquantel | 3.5 |
| Friis *et al.*, (31) | Zimbabwe | RCT | Children | 11 to 17 | Filtration | Praziquantel | 12 |
| Garba *et al.*, (22) | Niger | LC | Children | 6 to15 | Filtration | Praziquantel | 6 and 12 |
| Gyoten *et al.*, (32) | Kenya | BF (no control) | Community | All | Urinalysis | Praziquantel | 12 |
| Houmsou *et al.*, (33) | Nigeria | Follow-up | Children | 1 to 15 | Filtration | Praziquantel | 12 |
| Kabuyaya *et al.*, (34) | S. Africa | PC | SC | 10 to 15 | Filtration | Praziquantel | 5 and 7 |
| Lemos *et al.*, (35) | Angola | Longitudinal | Children | 2 to 15 | Centrifugation | Praziquantel | 6 |
| Mutsaka-makuvaza *et al.*, (36) | Zimbabwe | PC | PSAC | 0 to 5 | Filtration | Praziquantel | 12 |
| Mutapi *et al.*, (37) | Zimbabwe | BF (no control) | Children | 6 to 15 | Filtration | Praziquantel | 9 |
| Mduluza *et al.*, (38) | Zimbabwe | NA | Children | ≤5 | Filtration | Praziquantel | 24 |
| Ofoezie, (39) | Nigeria | NA | SC | 5 to 16 | Filtration | Praziquantel | 3, 6 and 12 |
| Senghor *et al.*, (40) | Senegal | LC | SC | 5 to 15 | Filtration | Praziquantel | 6 |
| Senghor *et al.*, (41) | Senegal | LC | Community | 5 to 60 | Filtration | Praziquantel | 7, 12 and 36 |
| Webster *et al.*, (42) | Senegal | NA | Children | 3 to 15 | Filtration | Praziquantel | 6 and 12 |
| *Schistosoma japonicum* |  |  |  |  |  |  |  |
| Belizario *et al.*, (43) | Philippines | RCT | SC | 10 to 19 | Kato katz | Praziquantel | 6 and 12 |
| Jiz *et al.*, (44) | Philippines | Cohort | Community | 7 to 30 | Kato-katz | Praziquantel | 6 and 12 |
| Li *et al.*, (45) | China | PC | Community | all | Kato-katz | Praziquantel | 24 |
| Olliaro *et al.*, (26) | Philippines | RCT | Children | 10 to 19 | Kato katz | Praziquantel | 6 and 12 |
| Zhaosong *et al.*, (46) | China | BF (no control) | Community | 3 to 67 | Kato-katz | Praziquantel | 9 |

Note: PC = prospective cohort, BF = before and after, RCT = randomised clinical trial, LC = longitudinal cohort, NA = not indicated, SC = school children and PSAC = preschool aged children

#### 3.3.2 Schistosomiasis reinfection rate

The mean Schistosomiasis reinfection was rate 36.1% (±23.3%). The lowest reinfection rate (1.2%) was recorded in a study conducted in Zimbabwe after follow-up time of 12 months. Participants were preschool aged children of less than 6 year old living in an endemic area. The study participants were cured from *Schistosoma haematobium* infection after being treated with multiple doses of 40 mg/kg of praziquantel (every 8 weeks for 2 years). The highest reinfection rate (85.7%) was reported in a study conducted in Angola after follow-up time of 6 months. In this study, participants were school aged children at the community aged between 2 and 15 years old. The study participants were cured from *Schistosoma haematobium* infection after being treated with a single dose of praziquantel (40 mg/kg).

#### 3.3.3 Meta-analysis/subgroup analysis

The meta-analysis included 54 datasets extracted from 29 studies to calculate the pooled estimate of the Schistosomiasis reinfection rate (Figure 2). The pooled estimate of rate of reinfection was 33.2% (95% CI, 26.5 – 40.5%). Heterogeneity tests indicated high degrees of between-studies variances (*I*^*2*^ = 97.0%, p □ 0.001, and *r*^2^ = 1.348). Due to the presence of high heterogeneity, subgroup analysis was performed to check for the important contributing factors. The following factors were assessed for their contribution to the variance; type of *Schistosoma* species, geographical area, participants age group, follow up time, study settings and sample size. Subgroup analysis results showed that the type of *Schistosoma* species and age groups of study participants were associated with the pooled reinfection rate. The amount of heterogeneity contributed by *Schistosoma* species was *r*^2^ = 20.2%, p = 0.01 and age group was *r*^*2*^ = 4.4%, p = 0.07. The multivariate subgroup analysis shows that heterogeneity due to the combination of these two factors was *r*^*2*^ = 21%, p = 0.002. The pooled schistosomiasis reinfection rates according to the type of *Schistosoma* species were 44.3% (95% CI, 34.2% – 54.9%, *I*^*2*^ = 97%) for *Schistosoma mansoni*, 41.8% (95% CI, 30.9% – 53.5%, *I*^*2*^ = 96%) for *Schistosoma japonicum* and 19.4% (95% CI, 12.3% – 29.2%, *I*^*2*^ = 95%) for *Schistosoma haematobium*. The pooled reinfection rates were 25.7% (95% CI, 17/0% – 36.9%, *I*^*2*^ = 95%) for participants aged less than 16 year old and 38.89% (95% CI, 30.17% – 48.38%, *I*^*2*^ = 95%) for participants of all ages.

**Figure 2.**
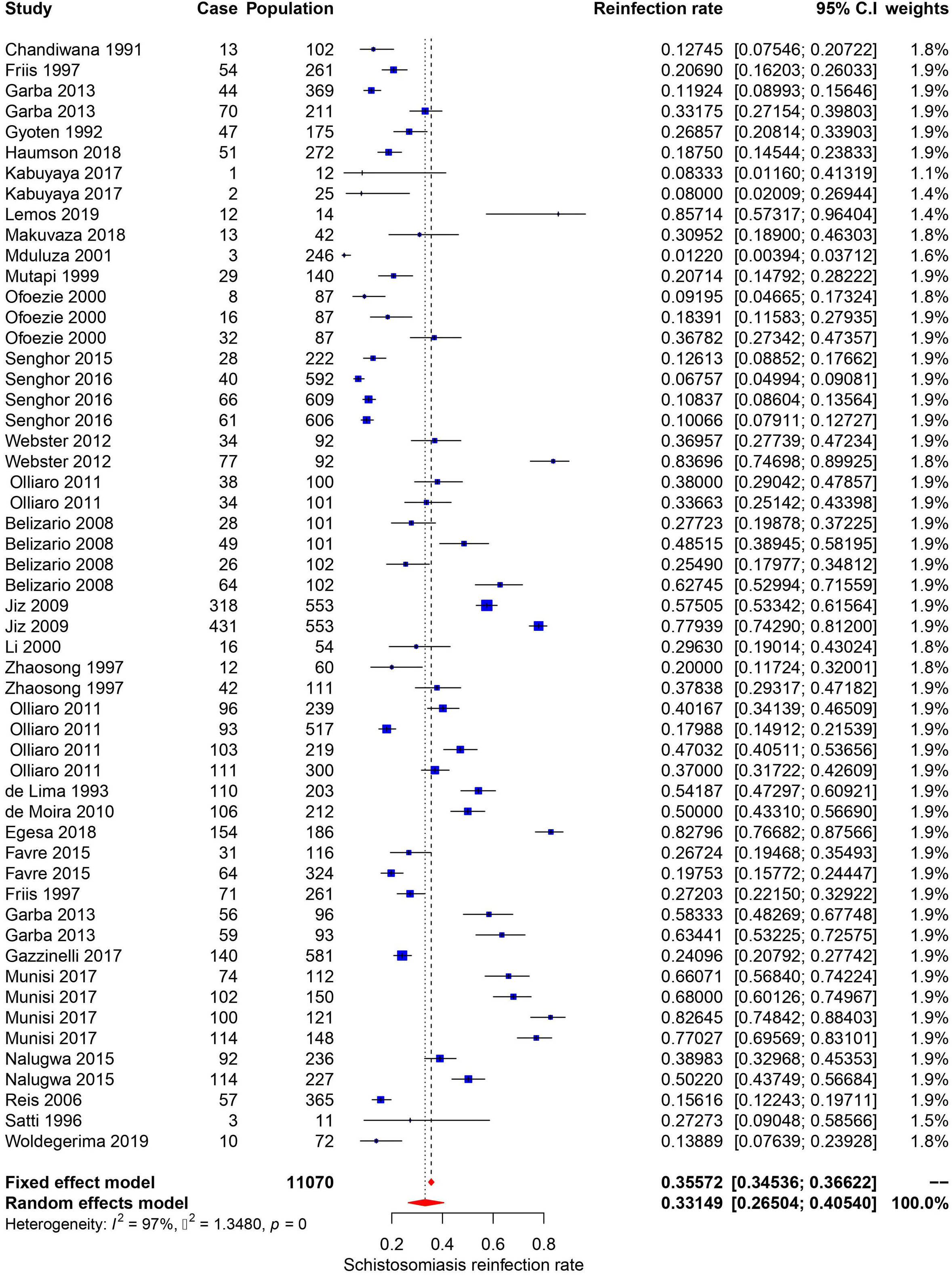
Forest plot of schistosomiasis reinfection rate

### 3.4 Intestinal schistosomiasis reinfection rate

#### 3.4.1 Studies characteristics

Intestinal schistosomiasis due to *Schistosoma mansoni* was reported in studies conducted in Africa (8 studies), Brazil (4 studies) and Africa and Brazil (1 study), while *Schistosoma japonicum* was reported in Philippines (3 studies) and China (2 studies). From 18 studies, a total of 32 datasets showing intestinal schistosomiasis reinfection rates were extracted. These datasets present reinfection rate after clients have been cured of the parasitic worm by praziquantel treatment (40 and/or 60 mg per kg body weight) as described in 17 studies and Oxamniquine (20 mg per kg body weight) as shown in 1 study. The study participants were people with age ranging from 1 to 67 years old. Ten studies were conducted at the communities, 7 studies were conducted at schools, and 2 studies were involved at both settings. All studies used Kato-katz technique for the identification and quantification of *Schistosoma mansoni* and *Schistosoma japonicum* eggs in stool samples. Shortest and longest follow-up times were 5 and 156 months reported in Tanzania and Brazil respectively (Table 1).

#### 3.4.2 Intestinal schistosomiasis reinfection rate

The mean reinfection rate was 43.9% (±20.6%). The lowest reinfection rate was recorded in a study conducted in Ethiopia after a follow-up time of 6 months. Participants were primary school children aged between 9 and 14 years. The study participants were cured from *Schistosoma mansoni* infection after being treated with a single dose of praziquantel (40 mg/kg). The highest reinfection rate was reported in a study conducted in Tanzania after follow-up time of 8 months. In this study, participants were school going children aged between 6 and 16 years leaving in an endemic area. The study participants were cured from *Schistosoma mansoni* infection after being treated with a single dose of praziquantel (40 mg/kg).

#### 3.4.3 Meta-analysis/subgroup analysis for intestinal schistosomiasis

The meta-analysis included 32 datasets extracted from 18 studies to calculate the pooled estimate of the intestinal schistosomiasis reinfection rate (Figure 3). The pooled estimate of reinfection rate was 43.4% (95% CI, 35.8 – 51.4%). Heterogeneity tests indicated high degrees of between-studies variances (*I*^*2*^ = 97.1%, p □ 0.001, and *r*^2^ = 0.836). Due to the presence of high heterogeneity, subgroup analysis was performed to check for the important contributing factors. Contributing factors analysed were type of *Schistosoma* species, age groups, study settings, follow up time, sample size and geographical area. Subgroup analysis results indicated that geographical area and age groups have an association with the pooled reinfection rate p = 0.02 and 0.09 respectively. The pooled intestinal schistosomiasis reinfection rates according to the geographic area was 53.7% (95% CI, 41.7% – 65.2%, *I*^*2*^ = 96.7%) for studies conducted in Africa, and 35.5% (95% CI, 27.5% – 44.4%, *I*^*2*^ = 96.5%) for studies conducted out of Africa. The pooled reinfection rates were 33.5% (95% CI, 22.6% – 46.4%, *I*^*2*^ = 96%) for participants aged less or equal to15 year old and 47.4% (95% CI, 38.2% – 56.7%, *I*^*2*^ = 97.2%) for participants of all ages (including below and above 15 years).

**Figure 3.**
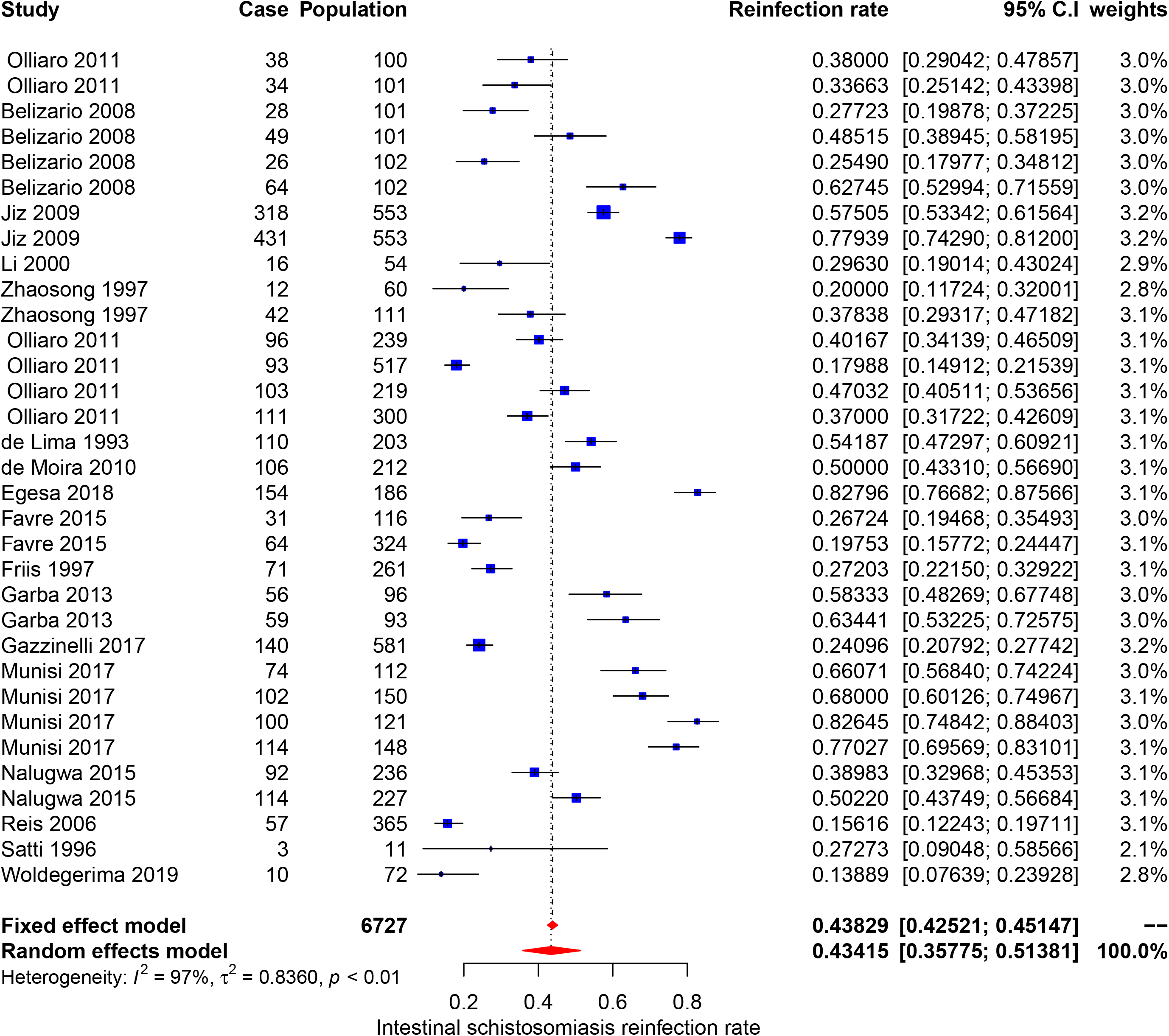
Forest plot of intestinal schistosomiasis reinfection rate

### 3.5 Urogenital schistosomiasis reinfection rate

#### 3.5.1 Studies characteristics

Urogenital schistosomiasis was reported in 14 studies conducted in Africa only (Table 1). From the 14 studies, a total of 22 datasets showing *Schistosoma haematobium* reinfection rate were extracted. These datasets present reinfection rate after clients have been cured from this parasitic worm by praziquantel treatment (40 mg per kg body weight). The study participants were people with age ranging from 2 to 60 years old. Eight studies were conducted at the community and 6 studies were conducted at school settings. Urine filtration (10 studies), centrifugation (1 study) and urinalysis (1 study) were the microscopic techniques employed for the identification and quantification of *Schistosoma haematobium* eggs in urine samples. Shortest and longest follow-up periods were 3 and 36 months reported in Brazil and Tanzania respectively.

#### 3.5.2 Urogenital schistosomiasis reinfection rate

The mean urogenital schistosomiasis reinfection rate was 17.6% (±10.8%). Generally urogenital schistosomiasis was detected at low and higher reinfection rate compared to intestinal schistosomiasis. Therefore, all information regarding urogenital schistosomiasis reinfection rates were provided in subsection 3.3.2.

#### 3.5.3 Meta-analysis/subgroup analysis for urogenital schistosomiasis

The meta-analysis included 22 datasets extracted from 14 studies to calculate the pooled estimate of the urogenital (*Schistosoma haematobium*) reinfection rate (Figure 4). The pooled estimate of reinfection rate was 19.4% (95% CI, 12.3% – 29.2%). Heterogeneity tests indicated high degrees of between-studies variances (*I*^*2*^ = 95.0%, p □ 0.001, and *r*^2^ = 1.4565). Due to the presence of high heterogeneity, subgroup analysis was performed to check for the important contributing factors. The assessed factors included age groups, study settings, follow up time, sample size and geographical area. Among the assessed factors, only sample size had an association with the observed heterogeneity. The amount of heterogeneity contributed by sample size was *r*^2^ = 25.6%, p = 0.014. The pooled urogenital schistosomiasis reinfection rates according to the sample size were 32.3% (95% CI, 14.9% – 56.5%, *I*^*2*^ = 95.2%) for studies with small sample sizes, 15.5% (95% CI, 9.3% – 24.5%, *I*^*2*^ = 95.3%) for studies with medium sample sizes and 9.2% (95% CI, 6.9% – 12.0%, *I*^*2*^ = 71.5%) for studies with large sample sizes.

**Figure 4.**
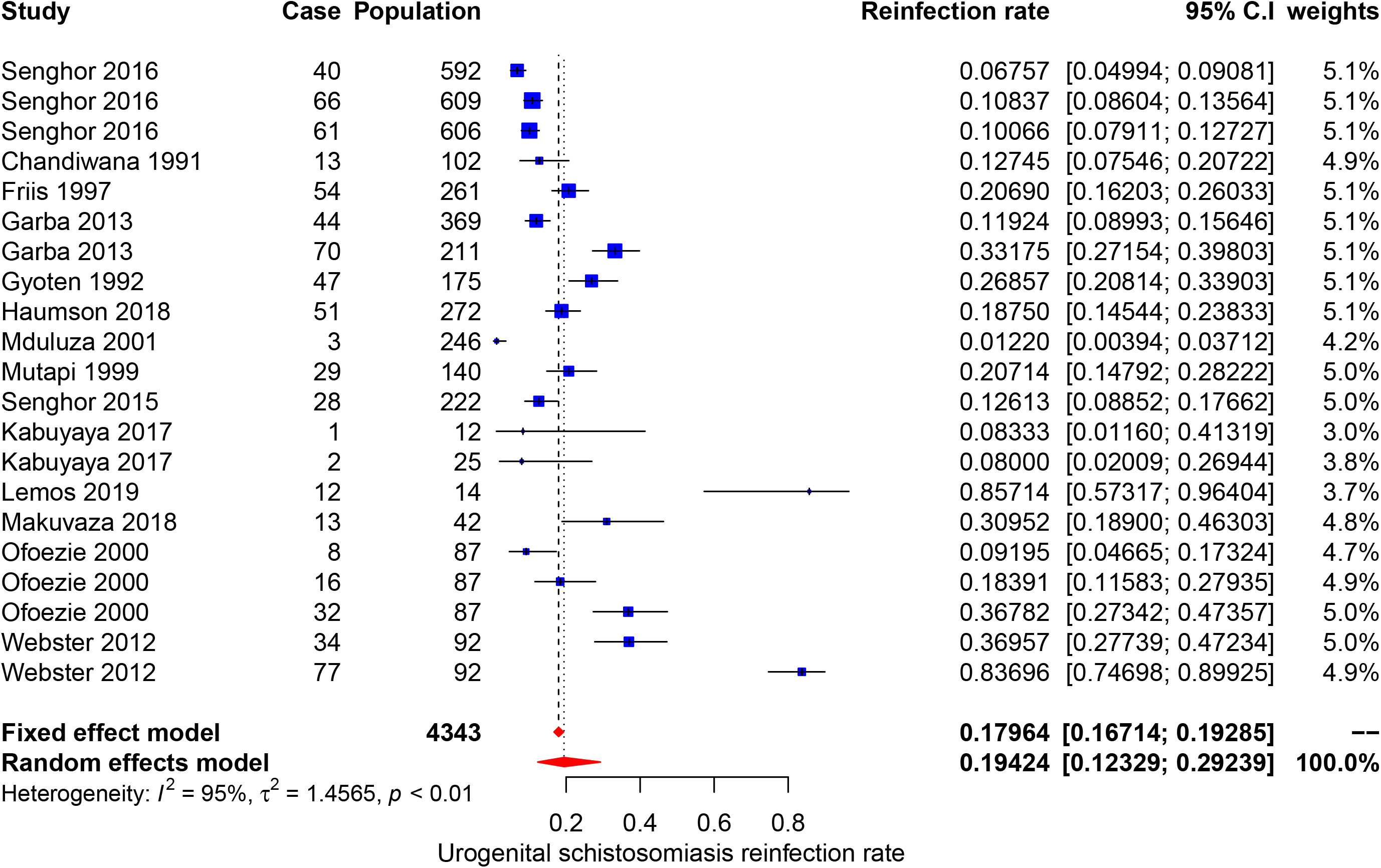
Forest plot of urogenital schistosomiasis reinfection rate

### 3.6 Publication bias

Figure 5 is a graphical presentation of studies and absence of publication bias. Egger’s regression analysis for funnel plot asymmetry results indicated the absence of significant level of publication bias (p = 0.483).

**Fig 5.**
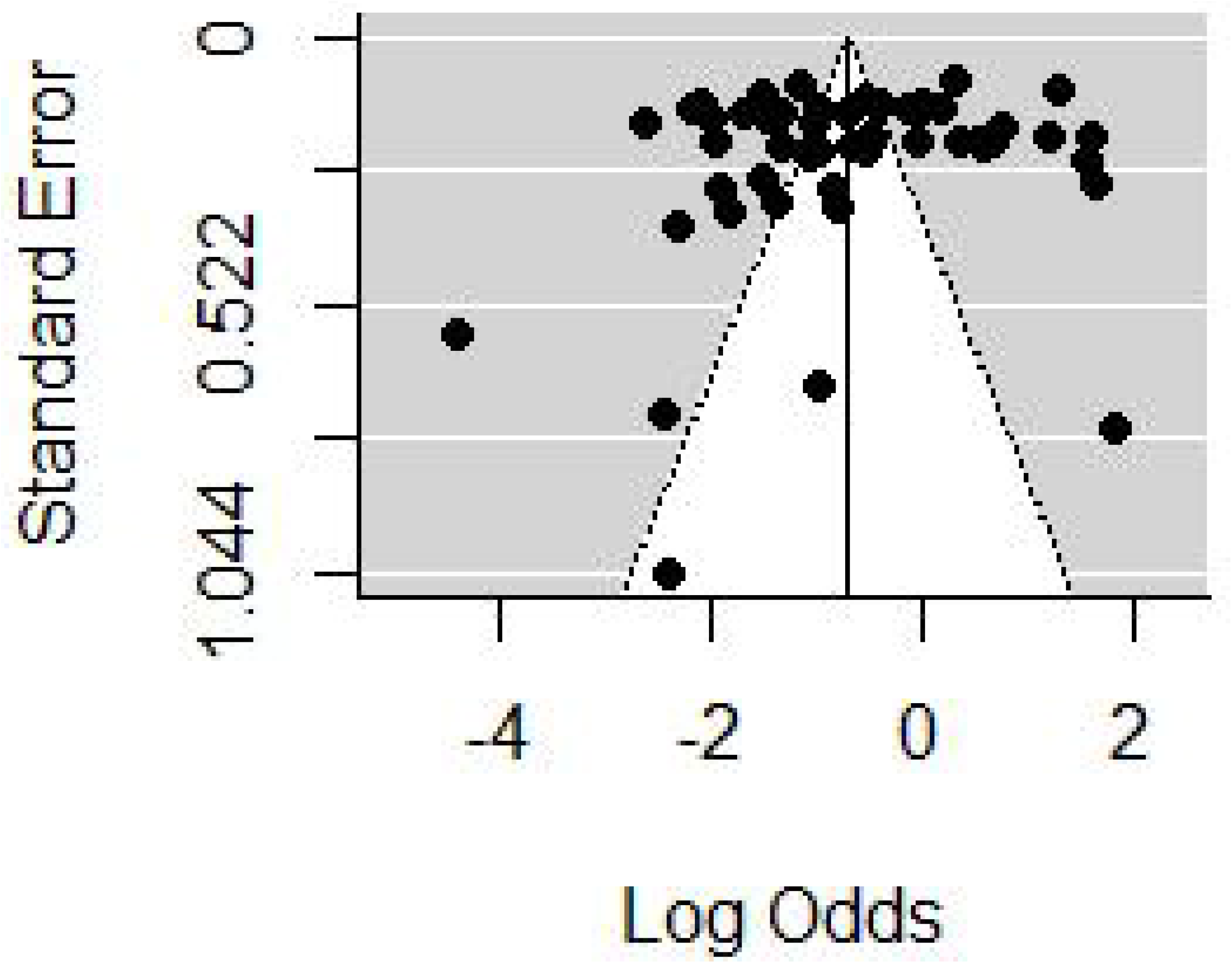
Funnel plot showing asymmetrical distribution for 29 (54 datasets) included studies

## 4 Discussion

Schistosomiasis is a serious human disease of public health concern, commonly presented in two forms namely intestinal and urogenital. To date the most successful means of controlling the disease is through chemotherapy by Praziquantel (43,47). In this review, we assessed the reinfection rate after administration of schistosomiasis treatment and the results are presented in terms of mean and pooled mean values. The mean value provides useful information at a population level to understand how many individuals were reinfected with *Schistosoma* species within a given population. Pooled mean takes into account the study size so that the contribution of each study to the meta-analysis reflects the number of individuals involved in each study. To our knowledge this is a first systematic and meta-analysis review that indicated the schistosomiasis reinfection rate of three predominant species globally following drug administrations.

The overall mean reinfection schistosomiasis rate was high (36.1%) with the minimum rate being 1.2% from Zimbabwe; as a result of intense repeated treatment in every eight weeks for a period of two years (38). The minimum value reported from Zimbabwe suggests that intensive repeated treatment of the disease for every two month for a period of 2-5 years will significantly control schistosomiasis infections. The observed pooled reinfection rate of 33.2% suggests that the reinfection rate is still higher worldwide amid the existing control programs in place hence, calling for effective operational strategies for the control and elimination of the disease.

The reinfection rate was further analyzed based on the two forms of schistosomiasis manifestation. The mean reinfection rate of intestinal schistosomiasis was 43.9%, lowest value being 13.9% from Ethiopia (29). Intestinal schistosomiasis had a pooled mean reinfection rate of 43.4%. Again the trivial differences between the calculated mean and pooled mean confirm the global high prevalence of intestinal schistosomiasis. The calculated mean reinfection urogenital schistosomiasis rate was 17.6% and pooled reinfection rate was 19.4%. The lowest reinfection rate of 1.2% from Zimbabwe was seen and is due to the factor explained above. Comparing the two forms of the disease urogenital schistosomiasis has the smallest reinfection rate comparing to intestinal schistosomiasis.

Several factors were found to influence high heterogeneity. Among six assessed factors the following contributed to the heterogeneity; type of *Schistosoma* species, age groups, sample size, geographical area and study settings. Generally the Schistosomiasis reinfection rate was contributed by the type of *Schistosoma* species and age groups. *Schistosoma mansoni* and *Schistosoma japonicum* had higher pooled reinfection rates than *Schistosoma haematobium*, therefore contributed more to the overall poo*led Schistosomiasis* reinfection rate (Table 2). Both *Schistosoma mansoni* and *Schistosoma japonicum* are transmitted from the human hosts to the environment (fresh water sources) through fecal contamination. High reinfection rate of *Schistosoma mansoni* and *Schistosoma japonicum* could be due to preferably defecation in or near fresh water sources because of privacy provided by vegetation growing surrounding the water sources and availability of water for washing after defecation. It was estimated that when a total of 991 people defecate near water source and go to clean in water, the amount of feacal particles deposited in is equivalent to amount of feaces contributed by 12 people who defecated directly in the water. Also, it was observed the eggs of *Schistosoma mansoni* and *Schistosoma japonicum* have high longevities than *Schistosoma haematobium* eggs (7). Pooled schistosomiasis reinfection rate of studies with participants aged less than 16 years were low compared to pooled estimates of studies with participants of all ages. This is unexpected results as we expected young children (less than 16 years old) to have high reinfection rate due to high rate of exposure and low protective immunity (15). Probably our findings could be due to fact that most of studies included in this meta-analysis involved adult populations who are occupationally exposed to high-risk of schistosomiasis infection (fishermen and rice paddy farmers). Additionally, 24 (77.4%) and 9 (39.1%) studies on intestinal schistosomiasis were conducted on participants of all ages and less than 16 years age group respectively. The high number of studies on intestinal schistosomiasis conducted on participants of all ages could be the reason for high reinfection rate in this subgroup compared to studies with participants aged less than 16 years as *Schistosoma* species causing intestinal schistosomiasis were observed to have higher reinfection rates.

The heterogeneity of intestinal schistosomiasis reinfection rate was contributed by age group and geographical area. Though, contribution due to geographical area was not statistically significant (p ≥ 0.1). It was observed that participants of all ages had higher reinfection rate compared to participants aged less than 16 years. This is because participants of all ages included adults who worked in routine occupations and domestic activities that exposed them to infested water. This is in contrast with other studies which showed children less than 16 years have higher reinfection rate compared to other ages (19). However, studies with all age groups included participants less than 16 years old. In the geographical area, studies conducted from Africa region had higher schistosomiasis reinfection rate compared to studies from non-African region. This could be due to different socio-economic status, climatic and environmental factors. Most studies included in this review were from low-income African countries where people are involved in high-risk water contact occupations compared to studies outside Africa. It has been observed that in low-income countries, people engage in agricultural activities, fishing, irrigation canal cleaning, car washing, gold prospecting and tin-mining which results in subsequent exposure to water hence increase the risk of reinfection (48). Also, climatic and environmental conditions play an important role in reinfection rate. Between the two identified intestinal *Schistosoma* species only *Schistosoma mansoni* is favoured with the climatic conditions of Africa region, while the climatic conditions of outside the Africa favour both survival of *Schistosoma mansoni* (South America) and *Schistosoma japonicum* (Asia). Based on our findings *Schistosoma mansoni* had high reinfection rate compared to *Schistosoma japonicum* and hence high reinfection rate in Africa region.

The variation in urogenital schistosomiasis reinfection rate was attributed by sample size. It was observed that the smaller the sample size the higher the urogenital schistosomiasis rate and vice versa. The reason for higher reinfection rate in studies with small samples could be due to the intensive follow up compared to studies with large sample size. Furthermore, the observed variation might be due to different level of endemicity of urogenital schistosomiasis. For example all data sets contributed to large sample size group were from a single study, therefore presented only a reinfection rate at single area.

We expected that study setting and follow-up time contribute on the pooled rate schistosomiasis reinfection (21,49). However, our findings revealed that there is no statistically significant association between these two factors and reinfection rate. Study setting was divided into two subgroups (school based and community) with the view that school based studies involved participants at higher risk than community based studies. Though this is not the case with our finding and might be due to inclusion of other high-risk groups in the community based studies.

Follow-up time was divided into two subgroups (< 12 months and ≥ 12 months). The findings showed that the difference in reinfection rates between the two follow up time subgroups was not statistically significant as opposed to our expectation (higher reinfection rates in studies with follow up time of ≥ 12 months). The results imply that reinfection occurs rapidly even before 12 months and remain steady.

## Conclusion and recommendation

For the successful elimination of human schistosomiasis and attaining 2030 health and wellbeing for all (sustainable development goal number 3), intercept of re-infection in rate is a core need. Global human schistosomiasis reinfection rate is still higher as stipulated in this review, with African countries being a hotbed for re-infection. This review also confirms that intestinal schistosomiasis have higher reinfection rate than urogenital. Re-infection rate can be triggered by different factors, hence the understanding the epidemiology patterns, biology of the parasite and social status in every society is essential. The existing control tool so far has not taken us to elimination goal. Is intensive repeated mass drug administration for a control of disease transmission feasible? That is yet still a myth. Swotting from Zimbabwe, this review suggests that, for the control of human schistosomiasis transmission biannual mass drug administration rounds for people of all age groups is essential, this strategy coupled up with robust snail control and health education campaigns will lead us to the elimination of schistosomiasis.

## Data Availability

The data used to support the findings of this study are available from the corresponding author upon request

## Acknowledgements

Authors would like to thank Ms Mary Joseph and Ms Eliza Lupenza for their assistance during review process.

## Author approval

All authors have seen and approved the manuscript

## Finding sources

The authors declare that no financial support was secured for this research work

